# Living with Long Covid: A Qualitative Analysis of Experiences, Coping Strategies and Care across the Illness Journey in Switzerland

**DOI:** 10.64898/2026.07.20.26358142

**Authors:** Rainer Böhm, Anja Frei, Christina Haag, Viktor von Wyl, Tobias Hoch, Dominik Menges, Thomas Radtke, Milo A Puhan, Felix Gille, Tala Ballouz

## Abstract

**Background:** Long Covid affects millions worldwide and disrupts the personal, professional, and social lives of those affected. Yet, insights on the day-to-day experience of living with Long Covid, how people adapt to the condition, and how they experience care remain limited.

**Methods:** Between November 2024 and February 2025, we recruited people living with Long Covid through Long Covid related studies and patient networks. Data were collected through a one-time semi-structured survey, completed using a speech-to-text feature with automatic transcription, and addressing 1) key events and experiences, 2) coping and support strategies, and 3) advice to others affected. We applied an inductive thematic analysis to develop a framework of key themes.

**Results:** We included 137 participants (median age 48 years, 73.7% women, median three years since SARS-CoV-2 infection). Analysis yielded 13 sub-themes within four key themes: medical issues; social, occupational and health care impact; barriers to recovery; and resources and strategies. Participants described a broad range of symptoms, most notably fatigue, cognitive difficulties, post-exertional crashes and psychological symptoms including depression and, in some cases, suicidal thoughts. These symptoms profoundly disrupted their social, working and family lives. In severe cases, independent living became impossible, with social isolation, severely reduced activity, and financial difficulties. Many described a long diagnostic journey in which symptoms were frequently dismissed as psychological and early advice to stay active that worsened their condition. The health care and social security systems were seen as ill-equipped to support people affected by Long Covid. With no effective causal therapies, treatment focused on symptom relief and participants tried many complementary and alternative treatments. Pacing was the only strategy widely used and perceived as effective in preventing crashes, alongside lifestyle adjustment, peer support, and maintaining hope.

**Conclusion:** These narrative accounts reveal the multidimensional burden of Long Covid, one that is exacerbated by how affected people are treated within the health and social systems. These findings underscore the need for empathic, knowledgeable care, validation of people’s experiences, and policy frameworks equipped to recognize and support people with Long Covid.

**Patient or Public Contribution:** This study is about the lived experiences of people affected by Long Covid. During the conceptualization phase, we consulted three people with lived experience of Long Covid to discuss the relevance of the research questions and study design. All participants received a newsletter with a plain language summary of interim findings. Following completion of this analysis, we conducted a focus group discussion with eight participants to validate our findings, identify gaps, and ensure that the findings accurately reflected their experiences. Feedback from this process informed the final manuscript.

## Background

A substantial proportion of individuals infected by SARS-CoV-2 develop Long Covid, or post COVID-19 condition as termed by the WHO, a multi-system condition characterized by persistent, often fluctuating symptoms (1,2). For many, its effects extend well beyond symptoms themselves, disrupting work, relationships, and daily life functioning, and exposing them to a health and social system that has been challenged by a condition that is not well understood (3). To date, most research has focused on symptom patterns, prevalence, and underlying pathophysiology (4), and a growing body of work has quantified its socioeconomic and occupational impact (5,6). Comparatively, less is known about how individuals experience and manage Long Covid in their daily lives.

Understanding how people experience and live with a chronic condition – their “illness journey”– offers insights that clinical and epidemiological studies alone cannot capture. Personal accounts reveal where needs go unmet and care falls short, and improve patient-centered care, empowering affected people to voice their experiences and feel heard and valued (7). They can also show how those affected manage and cope with the effects of a disease on day-to-day basis (8). As with many other chronic conditions, self-management is central to living with Long Covid. In the absence of established curative therapies, identifying what has helped patients may inform care for others and guide healthcare professionals in better addressing their needs.

Qualitative research on Long Covid has already identified recurrent themes. This includes, for example, protracted diagnostic and rehabilitation journeys, heavy burden of psychosocial challenges that significantly impact wellbeing and quality of life, and disruption to identity, work, and relationships (9–11). In their encounters with the healthcare systems, affected people have described a lack of empathy and understanding from healthcare professionals resulting in erosion of trust in the health system (12) and leaving many to rely on online resources and tools for information and symptom management (9,11). These studies have revealed important insights but share some limitations. Conducted within a framework of interviews or focus group discussions, they were constrained in their sample sizes. Some may have examined specific subpopulations such as those hospitalized, and most have primarily focused on describing how experiences unfold across a single domain (for example occupational or clinical), rather than across the breadth of a person’s life. Few have described how affected people cope and adapt over time. These voices, particularly those who have adapted to the condition, can play a direct role in shaping patient relevant care and support.

To address these gaps, we conducted the “Living with post COVID-19 symptoms” survey (13), a semi-structured survey in which participants were asked to tell their story living with Long Covid, via spoken responses captured through an automatic speech-to-text transcription. Inspired by an earlier “My Life with MS” study among persons living with multiple sclerosis (MS) (14), this approach allowed us to elicit narrative accounts of experiences across a large, varied sample. The survey aimed to capture three dimensions of Long Covid experience: 1) key events and challenges participants encountered across their health, personal, and professional lives, 2) coping strategies, support systems and resources they drew on, and 3) the advice they would offer to others affected.

## Methods

### Study design

This study adopted a qualitative research design, aiming to produce a detailed and comprehensive account of the experiences and perceptions of people living with Long Covid in Switzerland. The study was approved by the ethics committee of the Canton of Zurich (BASEC-Nr. 2024-01025) and we obtained electronic consent from all participants prior to participation. Reporting of this study was guided by the Standards for Reporting Qualitative Research (SRQR) (15).

### Participants and recruitment

We recruited participants between November 2024 and February 2025 through two main channels. The first was four pre-existing Long Covid related studies conducted at or in collaboration with the Epidemiology, Biostatistics and Prevention Institute (EBPI) at the University of Zurich (UZH): three observational studies that followed people who have been infected with or vaccinated against SARS-CoV-2 with regular evaluation of Long Covid (16–18), and one randomized placebo-controlled trial (PYCNOVID) of Pycnogenol® on patient-reported health status in people with Long Covid (19). The second channel comprised two Long Covid patient networks: Altea Long Covid Network and Long Covid Schweiz (20,21). Interested participants completed a screening questionnaire on REDCap electronic data capture tool (22,23) to determine eligibility. Participants were considered eligible if they were aged 18 years or older, reported symptoms related to Long Covid and/or had received a clinician-confirmed Long Covid diagnosis, and had good command of German. The study was restricted to German-speaking participants for feasibility purposes, as this is the largest linguistic region in Switzerland (24) .

### Data Collection

Data were collected via a one-time, semi-structured electronic survey on the Research Management Information System (RMIS), a secure digital health study platform developed by the Digital and Mobile Health Group at the UZH (25). Participants were able to complete the survey on a laptop or mobile phone.

The survey was structured around two main dimensions:

- The first dimension concerned key events and experiences since the start of the Long Covid symptoms, changes to daily routine, changes to professional and personal life, events related to worsening or improvement of symptoms, positive or negative experiences with the healthcare system, and periods of emotional challenges.
- The second dimension covered topics around coping and support strategies including support systems that participants found helpful, resources they felt were lacking from the health and social systems, and advice they would offer to others living with Long Covid.

In addition, participants completed questions on demographics, comorbidities, SARS-CoV-2 infection history and Long Covid onset, use of health services and medications, and impact of Long Covid on daily functioning (using the post COVID-19 functional scale PCFS (26)).

During the conceptualization phase, we consulted three people with lived experience of Long Covid to discuss the relevance of the research questions and study design. The survey was developed by the research team, informed by findings from the literature on lived experiences of Long Covid and built on a similar life-events survey previously conducted among people living with MS (14). The survey was tested and iteratively refined by members of the research team prior to its launch, including checks of question wording and functionality of the speech-to-text function.

The survey incorporated a speech-to-text function that automatically transcribed spoken responses into written text using Open AI Whisper Large-v3 (27), allowing participants to respond to questions conversationally and in their own words. Participants who preferred not to use the speech-to-text function could alternatively type in their responses (approximately a third of participants).

Responses were collected primarily in Swiss German (with automatic transcription into Standard High German) with a small number of participants responding in English. English responses occurred either due to participant preference, or because in very rare and early instances the system auto transcribed the responses into English, after which some participants continued the rest of the survey in English for consistency. Participants were able to review and edit the transcribed text before submitting it, providing an opportunity to correct any mistranscription of their own responses. To protect participant privacy, speech recordings were immediately transcribed and deleted automatically after transcription. All responses were read in full by native German speaking members of the research team and checked for potential transcription artifacts, grammatical errors, and implausible content that may arise from automatic transcription. Implausible content was deleted and obvious errors (e.g., repeated words, misspellings, or incorrect word transcriptions such as homophones) were corrected while maintaining participants’ original meaning.

### Data Analyses

All survey responses were imported to MAXQDA (VERBI Software GmbH), a qualitative data analysis software. We applied an inductive thematic approach (28) within a structured organizational framework for the four research questions: 1) key life events and experiences, 2) coping and support strategies, 3) perceived gaps in care, and 4) advice to others. Coding was conducted inductively, allowing themes and subthemes to emerge directly from the participants’ accounts rather being determined in advance.

Analysis followed three iterative stages, supported by three team meetings at key points in the process. Prior to coding, RB, TB, AF, and FG met to discuss the analytic approach, following which RB had a training in qualitative analysis and use of MAXQDA. In the first stage of the analysis, RB read and reread the data, noting initial and recurring ideas across and within the four major research questions. In the second stage, RB inductively coded a subset of survey responses (∼40 participants), grouping content by similarity of meaning. Codes within each domain were progressively grouped into broader categories based on conceptual similarity, from which a preliminary codebook was developed. The research team then met and discussed the preliminary codebook, examining the coding structure, definitions, and organization of codes. Based on the team’s feedback, RB refined and finalized the codebook. In the final stage, RB applied the final codebook to the full responses. Overarching key themes were refined through a final team meeting through which RB, TB, AF and FG reviewed the emerging themes and reached consensus on the final thematic structure.

Verbatim quotes representing the key themes were chosen and are reported to illustrate the main findings and to preserve the participants’ voices as closely as possible. English survey responses were translated to German, and coding was conducted in German. The coding and quotes included in this manuscript were translated to English using the Software DeepL and were checked by TB and RB for accuracy.

All participants received a newsletter with a plain language summary of interim findings and were invited to a focus group discussion. We received 23 responses of which 11 were invited to participate to reflect diversity of experiences based on age, sex, duration living with Long Covid, and impact on daily life activities. Following completion of this analysis, we held the online focus group discussion (11 June 2026), attended by eight of the 11 invited participants of the survey to validate our findings, identify gaps, and ensure that the findings accurately reflected their experiences. Feedback from this process informed the final manuscript.

## Results

Of 233 who consented to participate in the study, a total of 137 participants completed the survey and thus included in this analysis (Supplementary Figure 1). Median age was 48 years (IQR= 40 - 57) and 73.7% (n= 101) were women. Most participants (96.6%, n= 114) had a documented positive SARS-CoV-2 test, with a median time since infection of 3 years (IQR= 2.6 – 4.0). 80.7% (n= 96) had received a Long Covid diagnosis from a clinician. More than two-thirds (68.1%, n=81) reported at least moderate limitations to their daily life activities due to Long Covid-related symptoms and median self-rated health was 35 (out of 100) according to EQ-VAS (Table 1). Participants who completed the survey were slightly younger (48 versus 54 years) than those who did not and a higher proportion were women (73.7% versus 65.8%).

**Table 1.**
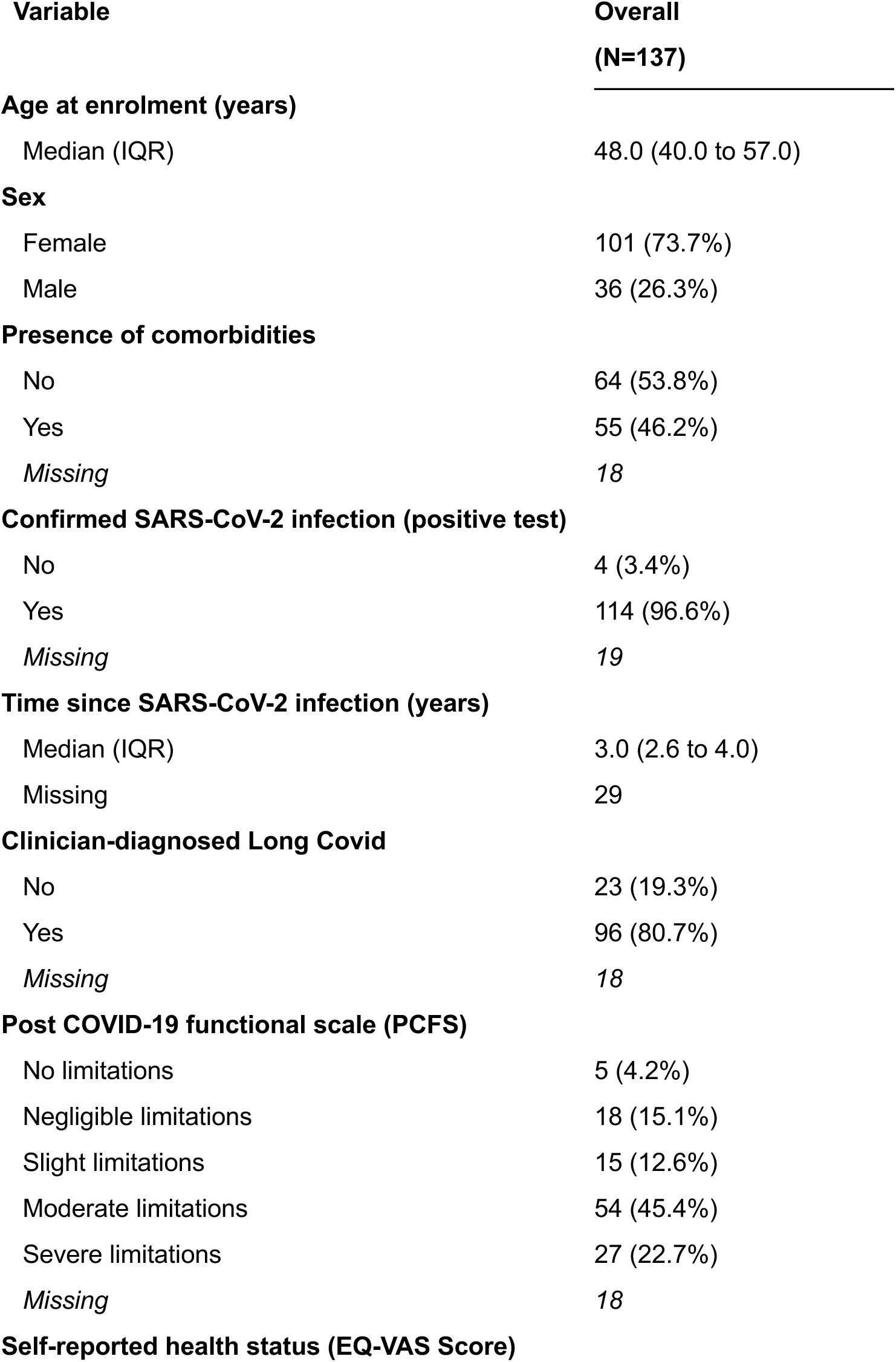
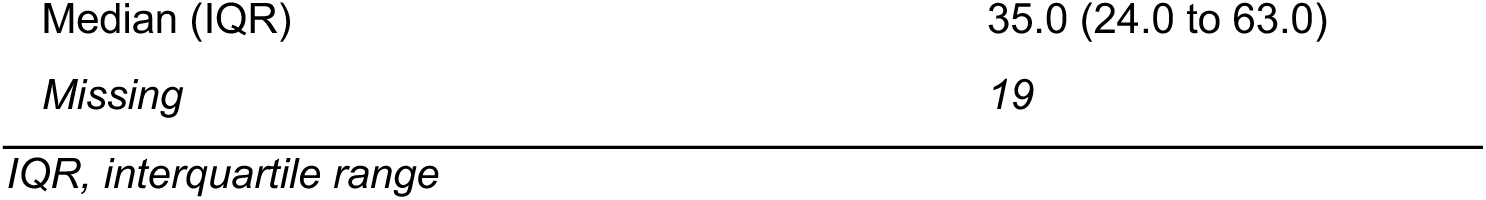
Characteristics of included study participants.

Through synthesis of a code system comprising more than 400 individual codes, we developed a framework of 13 sub-themes grouped into four key themes: medical issues, social, occupational and health care impact, barriers to recovery, and resources and strategies, that summarize participants’ experiences living with Long Covid (Table 2, Supplementary Table 1). Across these themes, participants’ accounts followed a broadly common trajectory from symptom onset and prolonged care journey, though episodes of worsening symptoms and crashes, to varying ways of coping and adaptation, and variable longer-term outcomes (Figure 1).

**Figure 1.**
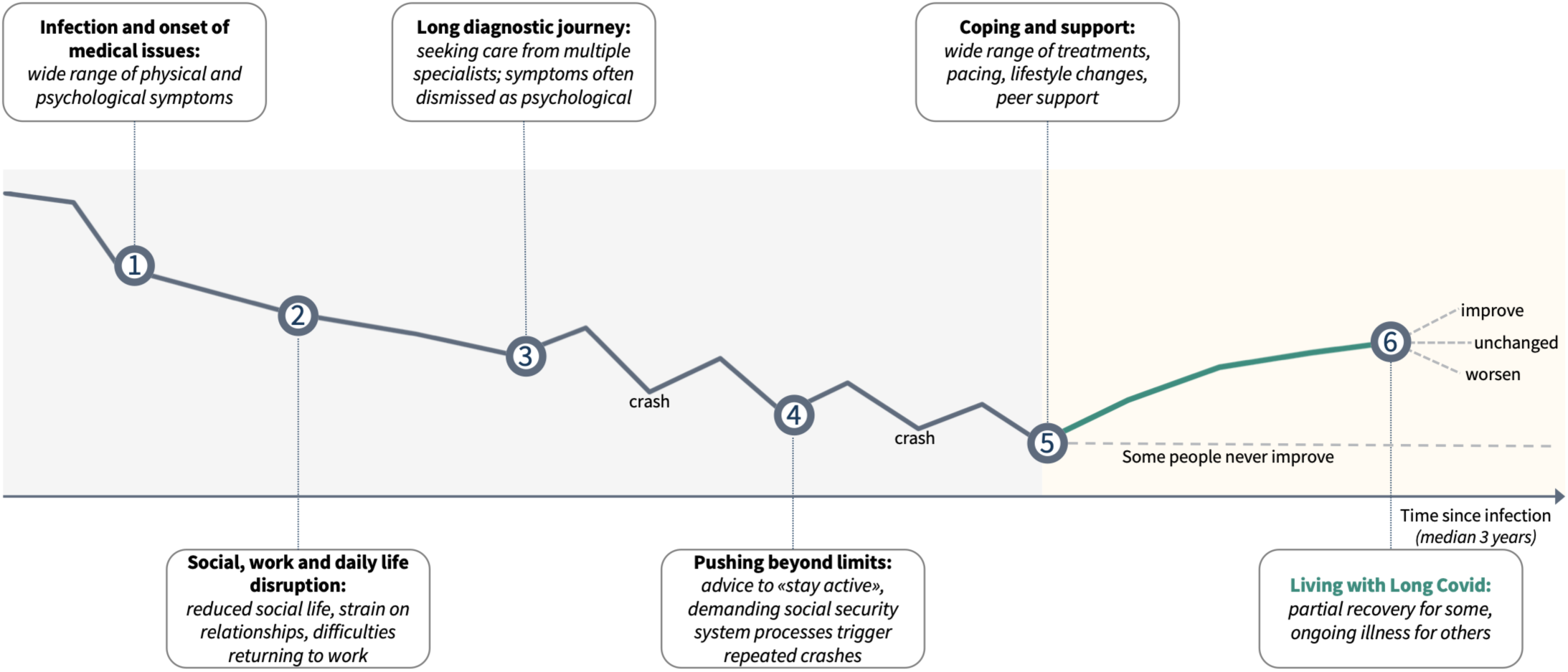
Illustrative Long Covid patient journey, synthesized from participants’ accounts and resulting themes

**Table 2.** Framework of themes identified and their reflected content.

| Key theme | Sub-theme | Explanation |
| --- | --- | --- |
| **Medical issues** | Range of Long Covid symptoms | Reflects the multitude and variety of symptoms |
|  | Psychological distress | Reflects the substantial mental and emotional challenges of the condition |
|  | Crashes | Reflects sudden and severe experiences of declines in physical and mental energy if personal limit is reached |
| **Social, occupational, and healthcare impact** | Disruption to social and work life | Reflects the profound impact on family, social, and working life and managing daily life |
|  | Experience with medical care | Reflects the long journey to diagnosis and care in the absence of a cure |
|  | Experience with social security system | Reflects interactions with systems not prepared for differing needs |
| **Barriers to recovery** | Pushing beyond one's limit | Reflects how over-exertion, self-driven or driven by early advice to stay active, worsens symptoms |
|  | Inadequate care | Reflects how a healthcare system grapples with an ill-defined condition |
| **Resources and strategies** | Health care professionals and treatments | Reflects the many health care professionals consulted and the conventional and alternative treatments tried |
|  | Pacing | Reflects the most broadly utilized approach to prevent and mitigate crashes |
|  | Lifestyle changes | Reflects how lives are being adjusted to cope with the condition |
|  | Optimism and hope | Reflects a mindset found helpful across many dimensions of illness and life |
|  | Patient groups and fora | Reflects the value of exchange, learning, mutual understanding and acceptance |

### Medical issues

Participants reported a broad **range of Long Covid symptoms** that could not be easily linked to identifiable organ damage, often contributing to a delayed diagnosis. Fatigue was experienced by almost all participants. Stress intolerance and a feeling of sensory overload were often combined with difficulties to concentrate and forgetfulness. Influenza-like symptoms, pain, breathing problems, tachycardia, and post-exertional malaise (PEM) were also reported. Together, these symptoms prevented participants from maintaining their former professional and social lives.

> “I was then at home for six months, only in my apartment. I felt physically very exhausted. I had difficulty concentrating, a kind of mental fog. I was very, very sensitive to noise” (Participant 1)

**Psychological distress** was frequent and manifested as emotional crises, fear, sadness, and depression. In a few instances, participants mentioned suicidal thoughts.

> “I couldn’t read; I couldn’t watch TV. I was just exhausted and, at the same time, feeling all torn up inside. All I wanted was to die. I cried; I wanted to die” (Participant 2)

Most participants experienced **crashes** as a result of being too active to maintain former routines or work, from overly strenuous rehabilitation or physiotherapy, and from sensory or emotional overload.

On reaching their individual limits, their capacity to function in daily life collapsed and they had to rest for days, often in dark rooms. Severe crashes became life-changing events and recovery was often slow, sometimes impossible.

> “I went for a walk around the house, but really just around the house – maybe 100 or 200 meters – and even that completely wiped me out, with nausea and everything. The symptoms became much worse. I was also much more sensitive to stimuli. And then, after about a week, the next crash came… And I got weaker every time” (Participant 3)

### Social, occupational, and healthcare impact

**Disruptions to social and work life** were severe and sometimes permanent. Participants described a much-reduced social life, and in severe Long Covid, extending to isolation and confinement at home. Daily activities and hobbies practiced prior to the infection were no longer possible. Managing daily life typically required help and was described as a turning point in their lives. While family and friends were described by many as a source of support, others reported a lack of understanding and few reported relationship breakdowns. Usual professional work was often not possible with frequent sick leaves and challenging attempts to return to work and sometimes leading to early retirement or disability status. Some experienced financial distress as a result of their condition and life became extremely difficult.

> “I have a chronic illness and am financially ruined because my health insurance provider, (…), refused to cover the cost of many treatments” (Participant 4)

The first point of **experience with medical care** was typically with a general practitioner (GP) with onward referrals to multiple specialists, who typically tested for and excluded organ damage.

Furthermore, prescribed therapies early on were often too strenuous and frequently led to crashes. In consequence, some participants left rehabilitation clinics in worse conditions than they had entered. While Long Covid clinics had been set up, care was often suboptimal due to lack of effective, causal therapies and due to lack of knowledge especially in the earlier years of the pandemic. Importantly, symptoms were often interpreted as psychological or psychosomatic in nature, a framing that fundamentally shaped participants’ clinical encounters and the treatments they received.

> “Conventional medicine couldn’t help me – that much was clear to me – apart from ruling out the possibility that there might be something else wrong with me” (Participant 5)

The **experience with social security system** was mixed; it was often described as ill-equipped to support the needs and reduced capacities of Long Covid participants. Its formal and demanding processes, requiring a series of hours-long interviews, appointments, forms, and medical examinations, were especially difficult for those with impaired cognitive and physical capabilities and often precipitated crashes. With no obvious pattern of organ damage being present, participants were sometimes labelled as fit for work and denied financial support.

> “I would like to see [disability insurance] assessors who understand the condition, who do not ridicule those who are unable to work, and who ultimately do not declare them 100% fit for work” (Participant 6)

### Barriers to recovery

Many participants described **pushing themselves beyond their limits** to keep up former levels of activities, especially in the early days of the pandemic when they were not aware of its harmful effects. This was aggravated by advice from healthcare professionals to stay active. Some had unrealistically high expectations of their abilities to cope with Long Covid and the speed of recovery, all of which leading to disappointment and ultimately crashes.

> “Looking back, I didn’t know anything about pacing back then and kept trying to push myself. But during those three months, I couldn’t do anything anymore – no TV, no books, no moving images. My body simply wouldn’t let me” (Participant 7)

**Inadequate care** manifested as lack of knowledge by health care providers on Long Covid and lack of empathy and understanding. Clinicians’ inability to help left participants feeling disappointed and alone. This was often paired with skepticism regarding the severity of their symptoms and participants felt dismissed. Combined with the lack of competent centers of excellence, participants felt being left alone to navigate through their disease journey and possible treatment options. After Long Covid clinics had been established, they received mixed reviews, depending on medical staff and affiliated institution. A lack of interest by politicians to learn about Long Covid and to take decisions to support research and better care was widely perceived.

> “Case management would have helped me; it would have shown me where to turn and what to do. Trying to organize everything on your own when you’re completely exhausted is very, very draining” (Participant 8)

### Resources and strategies

#### Health care professionals and treatments

In the absence of effective causal therapies, participants pursued a wide range of medical therapies primarily targeting symptom relief (Table 3, Supplementary Figures 2 and 3). They consulted many different specialists and tried a broad range of approved medications, experimental and off-label therapies, and complementary and alternative approaches. Approximately half (48.9%) had tried two or more classes of conventional/biomedical treatments and 32.1% had tried two or more alternative/complementary approaches. While some treatments offered partial symptom relief for some participants, no single approach was considered as widely effective.

**Table 3.** Healthcare professionals consulted and typical treatments tried by participants.

|  |  |
| --- | --- |
| Healthcare professionals consulted | General practitioners, physiotherapists, occupational therapists, pneumologists, cardiologists, psychologists, neurologists, psychiatrists |
| Conventional/biomedical treatments tried | Neuropathic/pain medications (30.7%), antihistamines (26.3%), immunomodulators (24.1%), antidepressants (18.2%), antiplatelet/antithrombotic medications (13.9%), antipsychotics (9.5%), antibiotics/antivirals, betablockers and autonomic heart rate medications (7.3% each), respiratory inhalers, medications targeting gastrointestinal system, metabolic/endocrine, and sleep/wake medications (6.6% each), corticosteroids (4.4%), and non-drug therapies such as apheresis (4.4%) and oxygen therapy (3.7%) |
| Complementary and alternative approaches | Dietary supplements (28.5%), herbal products (16.9%), meditation (10.2%), acupuncture (9.5%), traditional Chinese/Tibetan medicine (8.8%), breathing exercises (8.0%), osteopathy (6.6%), craniosacral therapy (6.6%), hypnotherapy (5.8%) |

*The list is not exhaustive, it reflects the most frequently reported professionals and treatment categories*

**Pacing** was the only reliable and broadly used strategy helping participants to cope with the condition. Being mindful of one’s own body, listening to warning signals, accepting the condition, and recognizing severely lowered individual limits were central to avoid crashes. Some participants used digital health trackers to identify what activities improved or worsened their condition. The ability to say no to oneself and others was seen as essential to maintain basic capabilities of managing daily life.

> “Keep calm, don’t overdo it, pace yourself. Anything else won’t do much good” (Participant 9)

Participants described **lifestyle changes** as necessary: slowing down, avoiding stressful situations and strenuous activities, limiting sensory input (for example with sunglasses or noise-cancelling earphones), using mobility aids like scooters or wheelchairs, and arranging domestic help were important. Practical and emotional support from a network of family and friends was seen as crucial, although this network became smaller and in a few cases the family situation deteriorated. At work, participants switched to part-time jobs or working from home, when this was possible. Formal reintegration into professional jobs was often attempted with mixed results; for some this was successful, others quit their jobs permanently, and some transitioned into early retirement.

> “It has helped me to adjust my lifestyle. I plan my day-to-day life quite differently now. I can only fit one appointment a day into my diary. As soon as there are several appointments, it can sometimes be too much” (Participant 10)

A mindset of **optimism and hope** went a long way to cope with the many limitations in daily lives and their psychological sequelae like depression. Maintaining hope, seeing the remaining positive things in life, appreciating them, and developing personal strategies for resilience was seen as important part of managing the life with Long Covid.

> “I think what helps me, too, is my positive and life-affirming nature, and my strong resilience. I find time and again that everything I’ve experienced in my life and invested in myself is now paying off” (Participant 11)

Joining **Long Covid patient groups and fora** provided participants with practical knowledge about the condition and possible treatments, helped them identify knowledgeable health care providers, and offered a space to share experiences. Beyond the informational value, peer exchange fostered a sense of not being alone on the complicated health care journey and provided emotional comfort and validation that participants missed in formal care.

> “I’ve been part of a Facebook group for people affected by Long Covid since 24 December, and I find it really supportive because there’s a wealth of knowledge there …. which gives me a sense of reassurance that I can actively draw on, and that helps me” (Participant 12)

Participants also described proactive efforts to build support around themselves: creating a Long Covid patient group when none were close by; systematically sharing study publications with their own physicians and therapists to improve referral and treatment decisions; and educating family and friends to foster understanding in their immediate social network. A few extended these efforts to politicians.

### Advice to others affected by Long Covid

When reflecting on what they would tell others living with Long Covid, participants’ advice mirrored the coping strategies mentioned above.

***Pace yourself and respect your limits*.** This was the most consistently given advice – to listen to one’s own body, recognize physical and cognitive limits, and slow down early.

***Connect with others who understand*.** Many advised seeking out and exchanging with others affected by Long Covid, through online fora or patient groups, not only for the sake of practical knowledge but also for finding comfort in not being alone in this journey.

***Inform those around you and accept help*.** Participants encouraged others to explain the condition to their network to foster understanding, and to seek and accept practical and psychological help rather than struggle alone.

***Hold on to hope and accept the situation*.** Participants advised others to accept this altered reality, notice the positive little things in life, and focus on what remains possible, and importantly never to lose hope.

***Find the right care*.** Participants highlighted the importance of finding a knowledgeable, empathic physician who takes them seriously and attending a specialized Long Covid clinic rather than relying solely on general practitioners. Many warned against physicians who did not take their symptoms seriously or dismissed them as “psychological” and urged “activation” which they regarded as harmful.

***Less is more*.** Advice regarding therapies, specifically off-label drugs, was heterogenous. While some encouraged others to try out different medications, dietary approaches, and psychological techniques, others warned against spending money and time on unproven therapies, cautioning that many of the therapies may have harmful side effects.

### Focus Group Discussion

The findings reported above were discussed during an online focus group with eight participants (7 females, 1 male, median age 48 years), all of whom had participated in the survey. Their illness course had since been stable, worsened or improved, exemplifying the heterogeneity in presentation of Long Covid. Overall, the group felt it was important to communicate the severity of Long Covid and its profound negative impact, at times amounting to the complete destruction of former lives. PEM and instant or delayed crashes were not only triggered by physical activities but also by strong emotional stress and often led to lasting deterioration in health. Dysautonomia was seen as central to many other symptoms while small fiber neuropathy was considered vastly underestimated and underdiagnosed.

Participants reported ongoing stigmatization, neglect and lack of understanding by physicians, media and the public. The social security system was still experienced as too demanding, with overburdening and inappropriate processes and little regard for the situations of those affected; the chances of disability being formally recognized were seen as very slim, and many had given up during the disability insurance process. Participants also highlighted physicians’ lack of knowledge, a tendency to place the responsibility for Long Covid down on patients themselves, and ongoing labelling as having psychological or psychosomatic problems. Competent centers of excellence and/or case managers were strongly missed.

Their key messages and demands were 1) urgent adjustment of evaluation processes within the social security system, 2) improved collaboration of Swiss health professionals with other countries who are investing more to support research and therapy, 3) inclusion of patient input in the design of future research, and 4) use of this study’s findings to raise awareness among the public and policy makers.

## Discussion

Our report reflects the *experiences* of individuals suffering from Long Covid and how they manage their daily lives – the condition seen through their own eyes instead of the eyes of doctors or caregivers. Our analysis of 137 participant narratives yielded four major themes being 1) medical issues, 2) social, occupational, and healthcare impact, 3) barriers to recovery and 4) resources and strategies. Together, these themes describe not only the enduring impact of Long Covid on the daily life of affected individuals but also the strategies they used to deal with it.

Assessed at around three years after SARS-CoV-2, participants reported a median self-rated health of 35 out of 100, and more than two third had at least moderate daily life limitations, highlighting how severely affected this sample was years after the infection. The symptom pattern they described, specifically fatigue, brain fog, and PEM that included recurrent severe crashes, is consistent with existing literature (e.g. (29,30)) and closely mirrors that of chronic fatigue syndrome ME/CFS (31,32). The recurrent crashes were central to the lived experience. They were perceived as severe and life-changing and often followed over-exertion, over-activating therapies or sensory or emotional overload. Many participants described how they had to learn the “hard way” to listen to the signals of their body and to respect their physical and emotional limits. Notably, advice from healthcare professionals early in the pandemic to stay active had led to such crashes, consistent with the post-exertional nature of the illness and the long-standing concerns regarding graded activity in such conditions (33). Together with evidence from cohorts showing that a substantial proportion of severely affected individuals do not recover (34,35), these experiences support an interpretation of Long Covid as a chronic post-viral condition that, in its severe form, becomes long-lasting and possibly permanent, as seen in Long SARS after SARS-CoV-1 infections (36–38).

Participation in family, social, and working life was disrupted with shrinking networks of family and friends, frequent sick leave and often unsuccessful or even harmful attempts at return-to-work leading some to go into early retirement or onto disability status. Anderson et al. describe this as “episodic disability” - those affected exist somewhere between being “able” and “disabled”, with sustained return-to-work unpredictable for both people living with Long Covid and employers (39). Standard “one-size-fits-all” return-to-work policies assume a stable recovery trajectory that those with Long Covid often do not have. In the absence of arrangements that are flexibly aligned with the condition, many engage in presenteeism, working despite symptoms and post-exertional crashes, or exit the workforce.

Participants’ experiences with medical care generally followed a long journey to diagnosis with referrals to multiple specialists to test and exclude organ damage. Symptoms were frequently treated as psychological; a pattern of “disregard and dismissal” that is now well recognized in Long Covid (40) and described more broadly as a form of epistemic injustice (41). This dismissal was perceived as actively harmful by participants. Being told that their symptoms were psychological led them to doubt their own experience which in turn drove many to ignore their symptoms and to push themselves to keep up with life and work, creating a vicious cycle of over-exertion and crashes. The psychological and emotional distress that participants described also needs to be understood in this light; not just as a consequence of the condition itself but also of the repeated experiences of invalidation.

Beyond dismissal, participants also reported a missing coordinating role or what Nguyen et al propose a “navigator role” in the Long Covid healthcare network (42). In the absence of a coordinated model of care, participants often had to organize their own care, moving between different specialists and sometimes being abandoned by them when no organ damage was found. Many felt that this was a result of lack of recognition of Long Covid as a physical condition and lack of clear diagnostic criteria. The few Long Covid clinics deemed by participants to have sufficient understanding of both the condition and patient needs were often overwhelmed, and many have since closed. This shifts Long Covid and more generally people with post-viral conditions to primary care without dedicated support, effectively leaving participants to act as their own care coordinators. Clinical guidance for primary care and frontline physicians exists (29,43). However, in the absence of integrated care, time and training on Long Covid, the impact of their implementation remains unclear.

Pacing was the only strategy that was consistently reported by participants to be beneficial, alongside accepting the condition and maintaining optimism and hope. Significant lifestyle changes were necessary to manage life without risking recurring crashes. To compensate for the lack of integrated care and knowledgeable physicians, participants consulted internet pages and Long Covid support groups. The exchange with other people affected by the condition helped them validate their experiences, created a sense of acceptance and allowed information exchange. Our findings on the value of such support groups align with Sarma et al 2025 (40), although they contrast with Milne et al 2025, where individuals previously hospitalized for COVID-19 reported found less comfort and validation from online support groups (44). This may reflect different needs after life-threatening acute illnesses. Nevertheless, this reliance on patient-led knowledge highlights the prominent role of these communities from the outset.

Treatment seeking was wide ranging. In the absence of specific, evidence-based drug therapies, participants consulted many specialists and tried a broad spectrum of medications including off-label and experimental drugs and complementary and alternative approaches. Many had tried multiple medications, reflecting both the many overlapping health challenges participants faced and the lack of guidance offered to them. For some, therapeutic decisions were informed by non-specialist sources, such as patient support groups, rather than by their physicians. This reliance on non-specialist information can be risky, as such sources are not always evidence based. It is understandable that individuals turn to these alternative sources when specialists are unable to help or dismiss their experiences, but this points to a need for better education and training of health care professionals in recognizing and managing Long Covid. This diversity of approaches also raises equity concerns. The ability to try multiple therapies, freely choose physicians and access additional medical and non-medical support may depend on individuals’ financial resources, risking a deepening of existing inequalities for those with fewer means.

Altogether, our findings point to several priorities. First, *knowledge and training*: health care professionals need better education on Long Covid, its symptoms and treatment options. Second, *care coordination*: centers of excellence are needed to coordinate the multi-specialty care that such a complex condition needs, with explicit navigator roles to reduce burden on patients. Third, *system adaptation*: social security and disability processes need to be redesigned for patients whose illness limits their capacity to engage with those processes. Fourth, *research investment*: sustained support for basic research and clinical studies is needed to better understand pathophysiology and enable more personalized treatment that goes beyond symptom relief. The recent cuts to the NIH-funded RECOVER initiative (45) make this priority more urgent. Interestingly, these priorities largely echo those identified through a citizen-driven, explicitly patient-centered research agenda set early in the pandemic (46). Effective treatment, coordinated care, improved awareness and knowledge among healthcare professionals, and support for daily and working life were among the top ranked priorities at that time (46). That similar priorities emerge from our study years later suggests that, despite increased attention to Long Covid, meaningful progress remains limited.

Several years after the pandemic, political and public attention has shifted elsewhere. Yet, waves of SARS-CoV-2 infections continue and will result in new Long Covid cases, albeit likely not in as high numbers as early during the pandemic. The patients most affected are the ones who are least able to advocate for themselves, with reduced energy and agency and potentially confined to a life at home. Ensuring that this group is not lost from sight, while also serving the larger group of people with milder, self-resolving symptoms is a public health and clinical priority that will remain for many years to come.

## Strengths and Limitations

Strengths of the study include the use of a speech-to-text survey, which allowed the inclusion of a comparatively large and varied sample size. This design lowered the participation burden for a population with limited energy and reduced capacities and likely enabled the inclusion of more severely affected participants than a written format would have. The semi-structured format gave participants more control over what they wanted to say, and open-ended questions allowed broad capture of their lived experiences beyond what we had included in the questionnaire design. Discussion of the results during the focus group discussion strengthened confidence in our findings.

Some limitations should be noted. First, despite the diversity of the sample, it was limited to German-speaking adults primarily in Switzerland which restricts the transferability of our findings to other geographical regions or healthcare and social contexts. Second, we included people if they had either a clinician-confirmed Long Covid diagnosis or self-reported symptoms consistent with Long Covid. While around 81% of included participants met the former criterion, the remaining were included based on self-report alone. Therefore, some participants in this self-report group may not have met diagnostic criteria or may have had symptoms attributable to other conditions, potentially affecting the comparability of experiences across the sample. This reflects a common challenge in Long Covid research given the absence of a validated diagnostic biomarker. Third, selection bias may have affected our sample. People who completed the survey were slightly younger and a higher proportion were women compared to those who did not. However, we could not assess whether these two groups differed in terms of Long Covid severity, digital literacy, or other relevant characteristics. People with higher digital literacy or familiarity with speech recordings may have been more motivated to participate. Patients with more severe symptoms may have been more motivated to participate and share their experiences or, conversely, may have participated less due to fatigue or if they found the survey cognitively too challenging. In a few instances, survey answers were extremely brief or stopped abruptly; a potential explanation being that those most severely affected could not sustain the concentration required by the survey. Fourth, recall bias may be a further concern as participants reflected on events spanning several years and forgetfulness is itself a typical Long Covid symptom. Finally, analyzer bias is an inherent trait of such qualitative analyses. We mitigated this through incremental development of the code system, coding of a subset of interviews by two other researchers and regular discussion of the results.

## Conclusions

Listening to the lived experiences of individuals with Long Covid reveals what clinical trials focusing on treatment strategies and observational studies focusing on prevalence and symptoms tend to miss – a chronic, ill-defined, and life-changing condition within health and social care systems that are not built to deal appropriately with it. In the absence of a system able to provide coordinated, multispecialty care over years, those affected become their own coordinators, developing their own strategies to compensate for the gaps. Yet, this self-reliance fails those who are severely affected without the capacity to advocate for themselves. Closing the gap will require knowledgeable clinicians, case managers and centers of excellence, and research that moves care beyond symptom-based support. Analyzing the experiences of affected individuals can guide such measures and importantly should be undertaken early when new diseases occur.

## Author Contributions

**Rainer Böhm**: methodology, investigation, formal analysis, writing – original draft, writing – review and editing. **Anja Frei**: methodology, supervision, project administration, writing – review and editing. **Christina Haag**: conceptualization, project administration, software, data curation, writing – review and editing. **Viktor von Wyl**: software, conceptualization, writing – review and editing. **Tobias Hoch**: project administration, writing – review and editing. **Dominik Menges**: project administration, writing – review and editing. **Thomas Radtke**: project administration, writing – review and editing. **Milo A Puhan**: conceptualization, writing – review and editing. **Felix Gille**: supervision, methodology, writing – review and editing. **Tala Ballouz**: conceptualization, project administration, methodology, data curation, supervision, validation, visualization, writing – original draft, writing – review and editing.

## Data Availability

The data that support the findings of this study are available upon request for researchers submitting a methodologically sound proposal to the corresponding author.

## Acknowledgements

The authors would like to thank the study participants for taking part in the survey and sharing their experiences as well as for their inputs in the focus group discussion. We would also like to thank Prof. Dr. Onur Boyman at the University of Zurich and the study administration team for their invaluable support.

## Ethics statement

The study was approved by the ethics committee of the Canton of Zurich (BASEC-Nr. 2024-01025) and all procedures were conducted in compliance with this approval.

## Consent

We obtained electronic consent from all study participants prior to participation.

