## Supplementary Material for "Living with Long Covid: A Qualitative Analysis of Experiences, Coping Strategies and Care across the Illness Journey in Switzerland"

#### Supplementary Methods

##### Rigour and reflexivity

TB, an infectious diseases physician and epidemiologist, is the principal investigator of the study. RB, a retired health care professional undertaking a Master's in Public Health during the study, led the analysis with input from TB, FG, and AF (public health professionals, social scientists, and epidemiologists). TB and AF have been actively involved in SARS-CoV-2 research generally and Long Covid research specifically through several studies conducted at the EBPI. Prior to this study, RB previously served as Chairman of the Board of a biotechnology company developing a treatment for Long Covid. This shared background in Long Covid research may have shaped familiarity with clinical framings of the condition. This was balanced by FG who has not been previously involved in Long Covid research thereby bringing an external perspective to the analysis, and by ongoing discussion of decisions and assumptions at team meetings during the coding and analysis processes.

As data were collected via a semi-structured speech-to-text survey rather than interviews, no researcher was present during data collection thereby mitigating interviewer-related confirmation and moderator biases. However, social desirability bias may still have shaped participants' responses as they were aware that their responses were being recorded and used for research purposes.

To further enhance rigour, findings were validated through an online focus group discission with a subset of participants (see Methods in main manuscript) to validate findings, identify gaps, and ensure that the final thematic structure accurately reflected participant' experiences.

### Supplementary Results

#### Supplementary Figure 1. Flowchart of participant enrolment

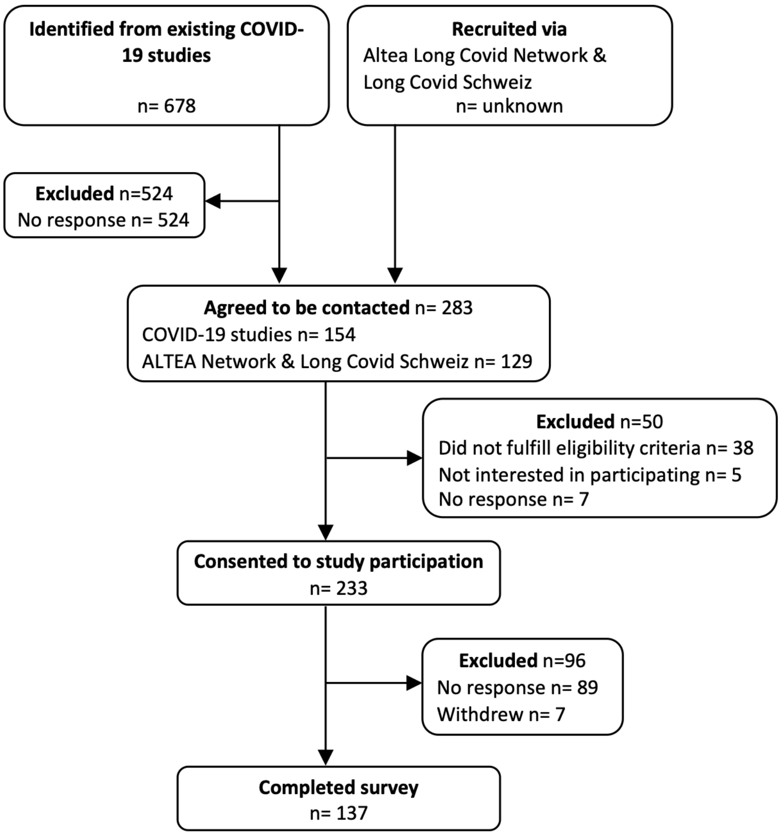

**Supplementary Table 1. Relationship of questions and codes. Categories of the code system were developed to reflect the full range of input given by the participants, even if not directly related to a research question; 1st-level codes group lower-level codes based upon similarity of content.**

| **Research Question** | **Survey Question** | **Category of Code System** | **1st-level Codes** |
|---|---|---|---|
| | Long Covid-related events | Course of (Long) Covid disease | SARS-CoV-2 Infection; Vaccination; Diagnosis; Development of Symptoms |
| | Long Covid-related events | Medical care utilized | GP; Multidisciplinary; Alternative; Outpatient; Inpatient; Study |
| | Long Covid-related events | Experience with medical care | Doctors; Therapists; Rehabilitation;<br>Long Covid Clinic; Hospital |
| Health-related challenges | Long Covid-related events | Health-related challenges | Crash; Fatigue; CVS & Lung; Mental Health & Central Nervous System; Gastrointestinal; Various Symptoms |
| Personal & social challenges | Long Covid-related events | Personal & social challenges | Personal; Environment |
| Professional challenges | Long Covid-related events | Professional challenges | At Work; regarding Social Security |
| How do daily lives differ | Long Covid-related events | Changes in daily life | Restrictions; Implications |
| How do patients adapt | Coping strategies | Which coping strategies were utilized | Pacing; Learning; Changes to daily Life; Support: Mental Health, Social Environment, Workplace, Social Security, Healthcare System |
| How do patients adapt | Helpful support | Which coping strategies were utilized | Pacing; Learning; Changes to daily Life; Support: Mental Health, Social Environment, Workplace, Social Security, Healthcare System |
| | | Which personal behaviour creates hurdles to improvements | Carrying on as if nothing were wrong; Refusing to acknowledge the Illness |
| Which support networks are being developed | Helpful support | Which networks & systems were developed | Awareness-raising; Self-help Groups; Networks within one's Personal Circle |
| Which gaps do patients identify | Gaps in public health | Which gaps in care were identified | Medical Care;<br>Long Covid Treatments; Social Security; Other Gaps |
| Which advice do patients give to other patients | Advice to other patients | Which advice is given to other patients | Making lifestyle Changes; Using the Healthcare System; Pacing; Information and Discussion; Other Advice |

**Supplementary Figure 2. Medications and non-drug interventions used as reported by participants to manage Long Covid, grouped by therapeutic category**

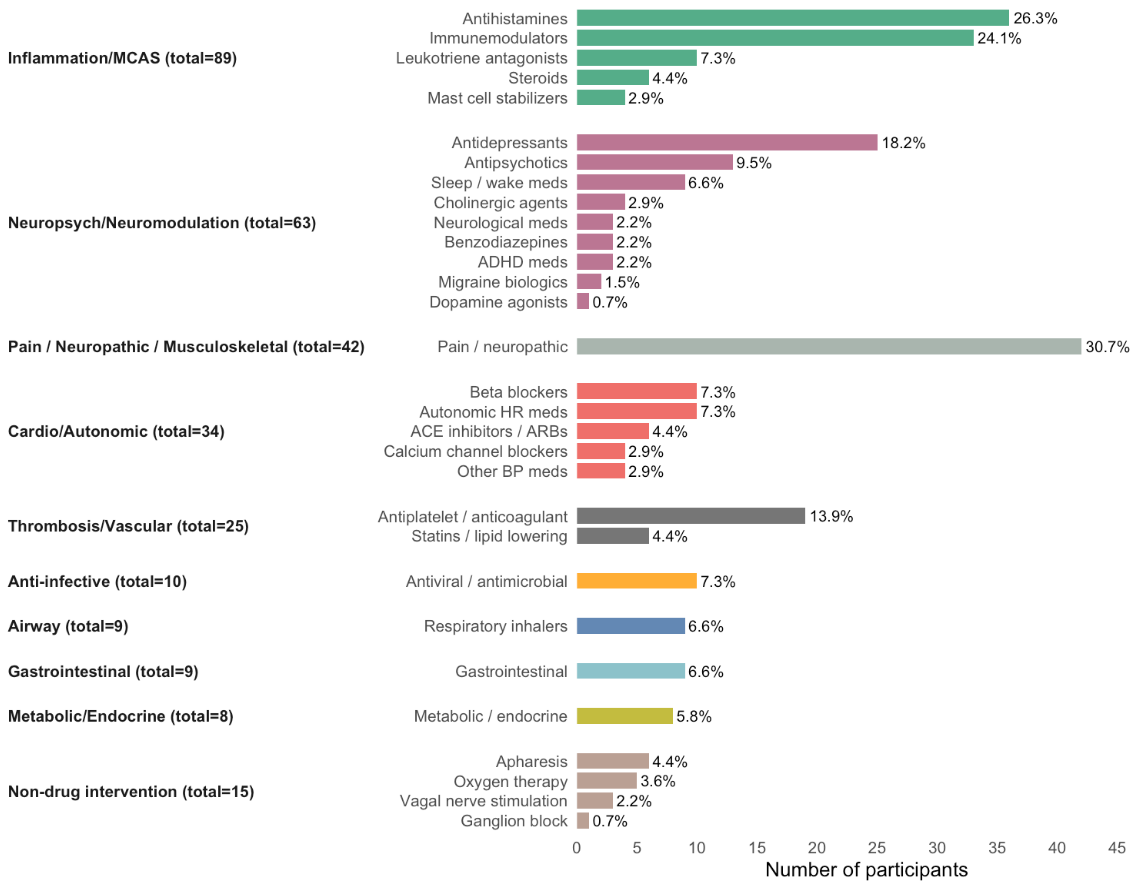

**Supplementary Figure 3. Complementary and alternative therapies used as reported by participants to manage Long Covid, grouped according to the NIH/NCCAM classification (1)**

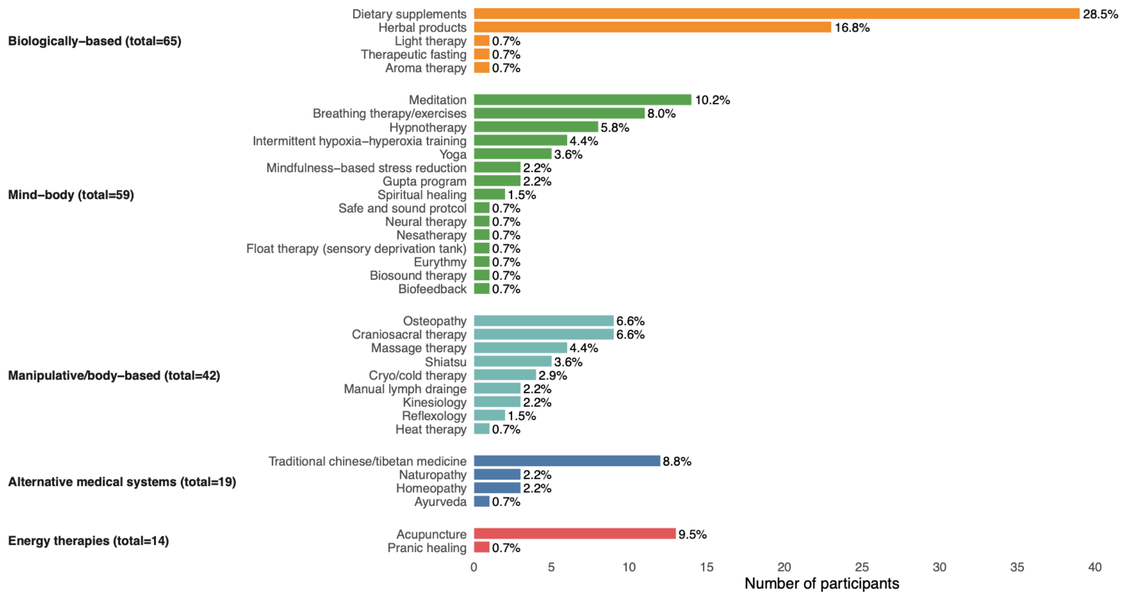

#### Out of the box Statements

While participants reported a broad range of symptoms and experiences, these were mostly mentioned several times. Here we report two improvements of symptoms that stood out of the other reports – thereby demonstrating the variety of (unexpected) experiences.

One female participant reported improvements of her Long Covid-symptoms during her pregnancy. One possible explanation might be the presence of autoantibodies that could have contributed to her form of Long Covid(2) and the known lesser immune response during pregnancy (Participant 13).

Another patient reported the disappearance of dizziness and clear thinking lasting for hours after a general anesthesia. This clarity of thought was experienced for the first time ever since the Long Covid diagnosis (Participant 14).
